# Nasopharyngeal metatranscriptomics reveals host-pathogen signatures of pediatric sinusitis

**DOI:** 10.1101/2024.03.03.24303663

**Authors:** Nooran AbuMazen, Vivian Chu, Manjot Hunjan, Briallen Lobb, Sojin Lee, Marcia Kurs-Lasky, John V. Williams, William MacDonald, Monika Johnson, Jeremy A. Hirota, Nader Shaikh, Andrew C. Doxey

**Affiliations:** Department of Biology, University of Waterloo, Waterloo, Ontario, Canada; Waterloo Centre for Microbial Research, University of Waterloo, Waterloo, Ontario, Canada; Cheriton School of Computer Science, Waterloo, Ontario, Canada; Firestone Institute for Respiratory Health, St. Joseph’s Hospital, Hamilton, Ontario, Canada; University of British Columbia, Department of Medicine, Vancouver, British Columbia, Canada; McMaster University, Department of Medicine, Hamilton, Ontario, Canada; University of Pittsburgh School of Medicine, Children’s Hospital of Pittsburgh of UPMC, Division of General Academic Pediatrics; Division of Infectious Diseases, University of Pittsburgh School of Medicine, Pittsburgh, Pennsylvania, USA

## Abstract

Acute sinusitis (AS) is the fifth leading cause of antibiotic prescriptions in children. Distinguishing bacterial AS from common viral upper respiratory infections in children is crucial to prevent unnecessary antibiotic use but is challenging with current diagnostic methods. Despite its speed and cost, untargeted RNA sequencing of clinical samples from children with suspected AS has the potential to overcome several limitations of other methods. However, the utility of sequencing-based approaches in analysis of AS has not been fully explored. Here, we performed RNA-seq of nasopharyngeal samples from 221 children with clinically diagnosed AS to characterize their pathogen and host-response profiles. Results from RNA-seq were compared with those obtained using culture for three common bacterial pathogens and qRT-PCR for 12 respiratory viruses. Metatranscriptomic pathogen detection showed high concordance with culture or qRT-PCR, showing 87%/81% sensitivity (sens) / specificity (spec) for detecting bacteria, and 86%/92% (sens/spec) for viruses, respectively. We also detected an additional 22 pathogens not tested for in the clinical panel, and identified plausible pathogens in 11/19 (58%) of cases where no organism was detected by culture or qRT-PCR. We assembled genomes of 205 viruses across the samples including novel strains of coronaviruses, respiratory syncytial virus (RSV), and enterovirus D68. By analyzing host gene expression, we identified host-response signatures that distinguished bacterial and viral infections and correlated with pathogen abundance. Ultimately, our study demonstrates the potential of untargeted metatranscriptomics for in depth analysis of the etiology of AS, comprehensive host-response profiling, and using these together to work towards optimized patient care.

**One Sentence Summary:** RNA-sequencing of nasopharyngeal samples enables pathogen-detection and host-response profiling in pediatric acute sinusitis.

## INTRODUCTION

Acute sinusitis is a bacterial superinfection that occurs usually in children with inflamed mucosa secondary to an upper respiratory tract viral infection (URTI) (*1, 2*). It is one of the most common diagnoses in pediatric primary care settings in the U.S. with 5 million antibiotic prescriptions written annually (*3*). However, because symptoms of acute sinusitis and an uncomplicated URTI overlap considerably, some children diagnosed and treated for acute sinusitis do not have a bacterial infection (*2, 3*). The diagnosis is especially challenging because the symptoms may be less specific in young children (*2*). Overtreatment of infections such as sinusitis is thought to be a major contributor to the rise in antimicrobial resistance (AMR), which remains an ongoing threat to public health (*1*).

Bacterial pathogens most frequently isolated from the sinuses of children with acute sinusitis include *Haemophilus influenzae* (HFLU), *Streptococcus pneumoniae* (SPN), and *Moraxella catarrhalis* (MCAT) (*2, 4*). Viruses such as influenza virus (INF), respiratory syncytial virus (RSV), coronavirus (COV), adenovirus (ADV), human rhinovirus (HRV), human metapneumovirus (MPV), enterovirus (EV), and parainfluenza virus (PIV) can produce symptoms that can be difficult to distinguish from acute bacterial sinusitis (*5*).

Recently, it has been suggested that one way to distinguish between bacterial and viral infections would be to obtain samples from the middle turbinate or nasopharynx of children with suspected sinusitis and to test these samples (using culture or qRT-PCR) for the three bacterial pathogens that frequently cause acute sinusitis (*6*). However, distinguishing bacterial sinusitis from an uncomplicated viral URTI using currently available microbiological tests is challenging for several reasons. First, pathogenic particles from asymptomatic infection or past infections may lead to false positive qRT-PCR detections that have little relevance to the presenting symptoms (*7*). Second, because many pathogens frequently colonize the nasal passages of children even during health, detecting their presence is often not indicative of the occurrence of a bacterial infection (*8, 9*).

With the remarkable reduction in the cost of high-throughput sequencing technologies, sequencing has emerged as an appealing strategy for the detection and taxonomic characterization of microorganisms in clinical samples from patients and has potential to overcome several limitations of currently available methods such as culture or qRT-PCR (*10, 11*). High-throughput sequencing of total RNA from a patient sample (metatranscriptomics) allows for a broad, untargeted approach to detect common, uncommon, and novel pathogens. Pathogen detection by high-throughput RNA or DNA-sequencing is showing promise in a growing number of infectious disease applications including pneumonia (*12, 13*), COVID-19 (*14*), meningitis (*15*), and febrile illness (*16*), and has been effective in identifying potential pathogens causing infection, including cases where no pathogen was detected using qRT-PCR or culture.

In addition, a significant benefit of metatranscriptomic sequencing is that it captures both pathogen-derived as well as host-derived RNA, which facilitates both pathogen detection as well as analysis of host gene expression patterns (host response profiling). Whereas sequence-based pathogen detection relies on detecting sequences of known pathogens, host-response profiling may quantify the expression level of biomarkers that indicate active host immune response to infection in a pathogen-agnostic manner. Thus, information on host-response may help distinguish active infections from colonization. Several previous studies have used RNA-seq or microarray techniques to identify and quantify biomarkers that differentiate between viral and bacterial respiratory infections (*17–22*). Using 104 host-response genes identified using microarray analysis of blood samples, Tsalik et al. developed separate bacterial and viral infection classifiers that had a combined accuracy of 87% (*17*). Host-response profiling from blood samples has also formed the basis of commercially available systems (e.g., MeMed BV®). If host-response profiles from a nasopharyngeal (NP) sample can similarly be used to differentiate bacterial from non-bacterial sinusitis infections, this could contribute to the development of biomarker assays that inform clinical decision making regarding the use of antibiotics.

In this work, to examine the ability of metatranscriptomics to uncover microbiological and clinically relevant information, we performed metatranscriptomic analysis of NP swabs from 221 children with clinically diagnosed acute sinusitis who were a subset of children enrolled in a previously described clinical trial (*6*). Through RNA-seq analysis of NP swab samples, we performed metatranscriptomic pathogen detection and assessed its ability to reproduce culture and qRT-PCR results for 3 bacteria and 12 viruses. We then assembled partial to complete genomes of 205 viruses. Finally, we performed host-response profiling and identified gene expression signatures of bacterial and viral infection, which correlated significantly with pathogen load. Our work shows the potential of metatranscriptomics for improving diagnosis of sinusitis and upper respiratory tract infections.

## RESULTS

### Cohort characteristics

A subset of 221 pediatric patients presenting with symptoms of acute sinusitis from a previous study (*6*) (Feb 2016 to Mar 2022) were selected for NP RNA-seq (Fig. 1, Table 1). Further details are provided in the Methods and in Shaikh et al. (*6*). One naris was sampled using a NP swab and this was used for viral qRT-PCR, bacterial culture, and RNA-sequencing (*23*); 171 (77 %) and 169 (76%) of the children tested positive for at least one bacteria or virus, respectively. Parents assessed symptom severity daily during the 10 days following diagnosis.

**Fig. 1.**
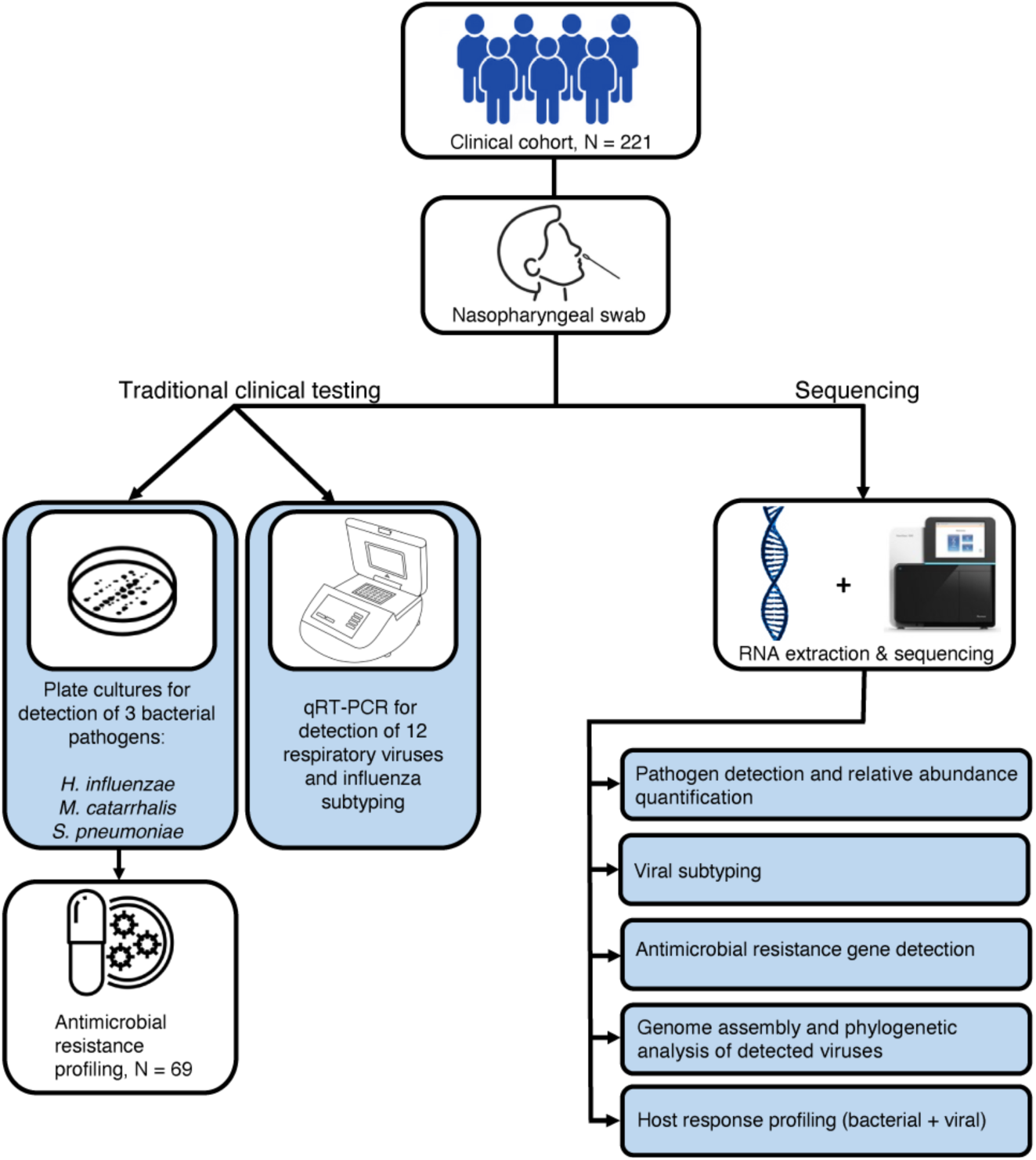
Overview of study design. The study cohort was comprised of 221 children with acute sinusitis who underwent collection of NP swabs. Culture was used to detect three bacterial species (*Haemophilus influenzae*, *Streptococcus pneumonia*, *Moraxella catarrhalis*) and qRT-PCR was used to detect 12 viruses of clinical relevance. *Haemophilus influenzae* isolates were tested for beta-lactamase production (N=69). Parallel to this, RNA extraction from NP swabs and sequencing was also done to conduct metatranscriptomic analysis using a bioinformatics approach. Using the sequencing data, several analyses were performed: pathogen detection and quantification, assembly of detected respiratory viruses, detection of beta-lactamase genes, and transcriptomic analysis of host responses.

**Table 1.** Demographic and clinical characteristics of pediatric patient participants with sinusitis. Demographic and clinical data for study cohort comprised of 221 children with persistent or worsening symptoms consistent with a diagnosis of acute sinusitis. Enrolled patients were assessed for symptoms and symptom severity. Pathogen detection for 3 common bacteria and a panel of 14 viruses was accomplished using culture and qRT-PCR, respectively.

| **Demographics** |  |
| --- | --- |
| Age (years)* | 4.8 (3.3–6.4) |
| Gender |  |
| Male | 115 |
| Female | 106 |
| **Clinical Characteristics** |  |
| Number of Days with Symptoms* | 14 (9–16) |
| Fever at any time during the illness | 121 |
| History of Asthma | 39 |
| History of allergic rhinitis | 64 |
| Coloured Nasal Discharge | 148 |
| **Clinical lab test results** |  |
| One or more bacteria detected | 171 |
| One or more viruses detected | 169 |
| Positive for beta-lactamase<sup>‡</sup> | 27 |

\* Median (interquartile range), <sup>‡</sup> Only samples positive for Hflu were tested (N = 69)

### Bacterial pathogen detection by metatranscriptomic analysis of NP samples

To identify potential bacterial and viral pathogens in the 221 samples, we performed high-throughput sequencing of total RNA derived from NP swabs. First, we aimed to quantify the abundance of three bacterial pathogens of interest –– *S. pneumoniae* (SPN), *M. catarrhalis* (MCAT), and *H. influenzae* (HFLU) –– as these pathogens are commonly isolated in children with bacterial sinusitis^4^. We note that our use of the term “pathogen” does not imply that these organisms are necessarily the causative agents of sinusitis infections. After quality filtering, we performed taxonomic classification of the sequencing reads using Kraken 2 (*24*). The relative abundance of the three bacterial pathogens (shown in Fig. 2A) was calculated based on the normalized abundance of reads (reads per million, RPM) that mapped to each species. One or more of these three bacterial pathogens were detected in a total of 177 patients (80%). Two or more bacterial pathogens were detected in 89 (40%) patients, and 25 (11%) of patients had all three bacterial pathogens detected. On an individual basis, SPN was detected in 73 (33%), MCAT in 137 (62%), and HFLU in 81 (37%) of patient samples. Tables S1 and S2 contain the clinical culture and RNA-seq based results for bacterial detection for each patient.

**Fig. 2.**
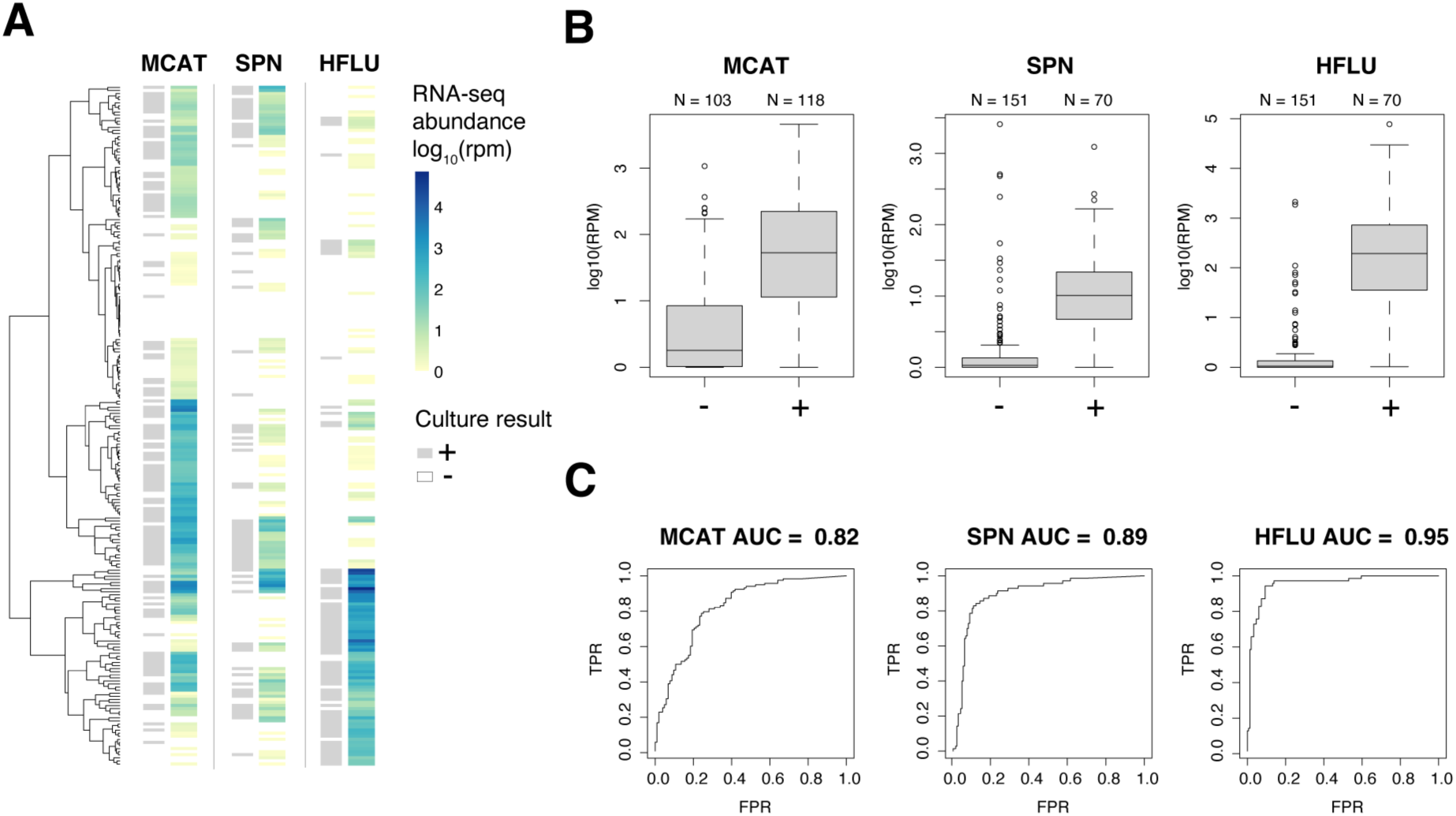
Metatranscriptomic detection of bacterial pathogens in NP samples from children with clinically diagnosed acute sinusitis. (**A**) Heatmap showing the detected abundance of three bacterial pathogens (*H. influenzae*, *M. catarrhalis*, *S. pneumoniae*) in patient metatranscriptomes. For each bacterium, the culture-based test result (positive – grey, negative – white) is shown on the left of the column, and the estimated RNA-seq abundance is depicted on the right of the column as a color gradient (absent – white, low – yellow, high – dark blue). Each row in the heatmap and tip in the hierarchical tree corresponds to an individual patient sample. (**B**) Boxplots depicting pathogen abundance in positive (+) versus negative (−) samples (labeled on X axis) defined based on culture. The boxes show the interquartile range and median line, and the whiskers show the variability extending to the furthest data points within 1.5 times above and below the interquartile range. Outliers outside of these ranges are shown as data points. (**C**) ROC curves illustrating specificity and sensitivity of metatranscriptomic pathogen detection with area under the curve (AUC) values displayed above.

Next, we examined the extent that the calculated abundance of these bacterial pathogens from RNA-seq agreed with their presence/absence based on culture. For all three pathogens, we detected a significant increase in RNA-seq abundance in those with a positive culture, demonstrating concordance between the metatranscriptomic data and culture (Fig. 2B). Some pathogen-negative samples based on culture had an RNA-seq pathogen abundance greater or equal to the mean abundance seen in positive samples. We then assessed the ability of the RNA-seq data to predict the culture-based test results for each pathogen, and generated Receiver Operator Curves (ROCs) by varying the detection threshold (Fig. 2C). HFLU infections could be detected with the highest accuracy by RNA-seq with an area under the ROC curve (AUC) of 0.95, SPN infections with an AUC of 0.89, and MCAT infections with an AUC of 0.82. Using a threshold of 3 reads per million, HFLU was detected with a sens/spec of 94%/90%, SPN with 81%/89% and MCAT with 85%/64% (Table 2).

**Table 2.** Sensitivity and specificity metatranscriptomics for detection of bacteria identified by culture or viruses identified by qRT-PCR.

|  | Sensitivity (%) | Specificity (%) |
| --- | --- | --- |
| **Bacteria** |  |  |
| *Moraxella catarrhalis* (MCAT) | 85 | 64 |
| *Streptococcus pneumoniae* (SPN) | 81 | 89 |
| *Haemophilus influenzae* (HFLU) | 94 | 90 |
| **Viruses** |  |  |
| Influenza A (INFA) | 100 | 94 |
| Influenza B (INFB) | 100 | 97 |
| Influenza C (INFC) | 33 | 96 |
| Human metapneumovirus (MPV) | 100 | 91 |
| Respiratory syncytial virus (RSV) | 90 | 92 |
| Human rhinovirus (HRV) | 73 | 77 |
| Parainfluenza virus 1, Human<br>respirovirus 1 (PIV1) | 100 | 94 |
| Parainfluenza virus 2, Human<br>orthorubulavirus 2 (PIV2) | 100 | 99 |
| Parainfluenza virus 3, Human<br>respirovirus 3 (PIV3) | 100 | 91 |
| Parainfluenza virus 4, Human<br>orthorubulavirus 4 (PIV4) | 91 | 91 |
| Adenovirus (ADV) | 44 | 97 |
| Enterovirus D68 (EVD68) | 100 | 90 |

### Beta-lactamase gene detection in HFLU positive samples

We next examined whether metatranscriptomics could identify potential resistance genes associated with HFLU. Culture-based tests for beta-lactamase were performed for all HFLU-positive samples, and these were used as the reference standard to analyze the accuracy of RNA-seq based detection. We assembled all non-human reads from samples that were clinically positive for HFLU (N=69) and used the Comprehensive Antibiotic Resistance Database (CARD) (*25*) to detect beta-lactamase genes with at least 10% coverage (fig. S1). Beta-lactamase genes were detected in 74% (20/27) of the samples associated with resistant HFLU, and in 33% (13/42) of the samples associated with non-resistance HFLU, which reflects a significant (2.1-fold) increase in detected beta-lactamase genes in the resistant samples (*p* = 0.002, Fisher exact test). The imperfect concordance between RNA-seq based and culture-based beta-lactamase detection reflects the known challenges in detecting AMR genes using metagenomic approaches (*26*). The complete list of genes and the portion of the reference genome detected for each hit can be found in tables S3-S5.

### Metatranscriptomic detection and analysis of respiratory viruses

To examine the ability of metatranscriptomics to detect viral infections, we first focused on respiratory viruses identified using qRT-PCR. Viruses tested for included influenza A (INFA), influenza B (INFB), influenza C (INFC), human metapneumovirus (MPV), human rhinovirus (HRV, which tested for rhinovirus types A, B, and C), parainfluenza virus 1 (PIV1), parainfluenza virus 2 (PIV2), parainfluenza virus 3 (PIV3), parainfluenza virus 4 (PIV4), respiratory syncytial virus (RSV, types A and B), human adenovirus (ADV), and enterovirus D68 (EVD68). One or more viruses were detected by metatranscriptomics in 175 patients (79%), two or more in 101 patients (46%), and three or more in 36 patients (16%). HRV was detected most frequently (45%), followed by MPV (14%) and INFA (13%).

Next, we examined the extent that the RNA-seq based predictions matched viral presence/absence based on the qRT-PCR. As shown visually in Fig. 3A, the relative abundance of viruses detected by metatranscriptomics was in strong agreement with the results of qRT-PCR-based tests, with lower qRT-PCR cycle threshold (Ct) values corresponding to higher RPM values in RNA-seq. A significant correlation (*r* = 0.75, *p* = 1.3x 10^-46^) was detected between 1/Ct values and viral load calculated as log_10_(reads per kilobase million, rpkm) (*27*) (Fig. 3B). Samples containing viruses detected by qRT-PCR but not by RNA-seq had significantly higher cycle thresholds (mean = 34.7) compared to true positives (mean = 23.2; t-test *p*-value = 5.5 x 10^-5^), which has been reported in previous RNA-seq studies (*28*). For all viruses except for INFC (which only had 8 positive samples), we detected an increase in metatranscriptomic abundance in those with a positive qRT-PCR result (Fig. 3C).

**Fig. 3.**
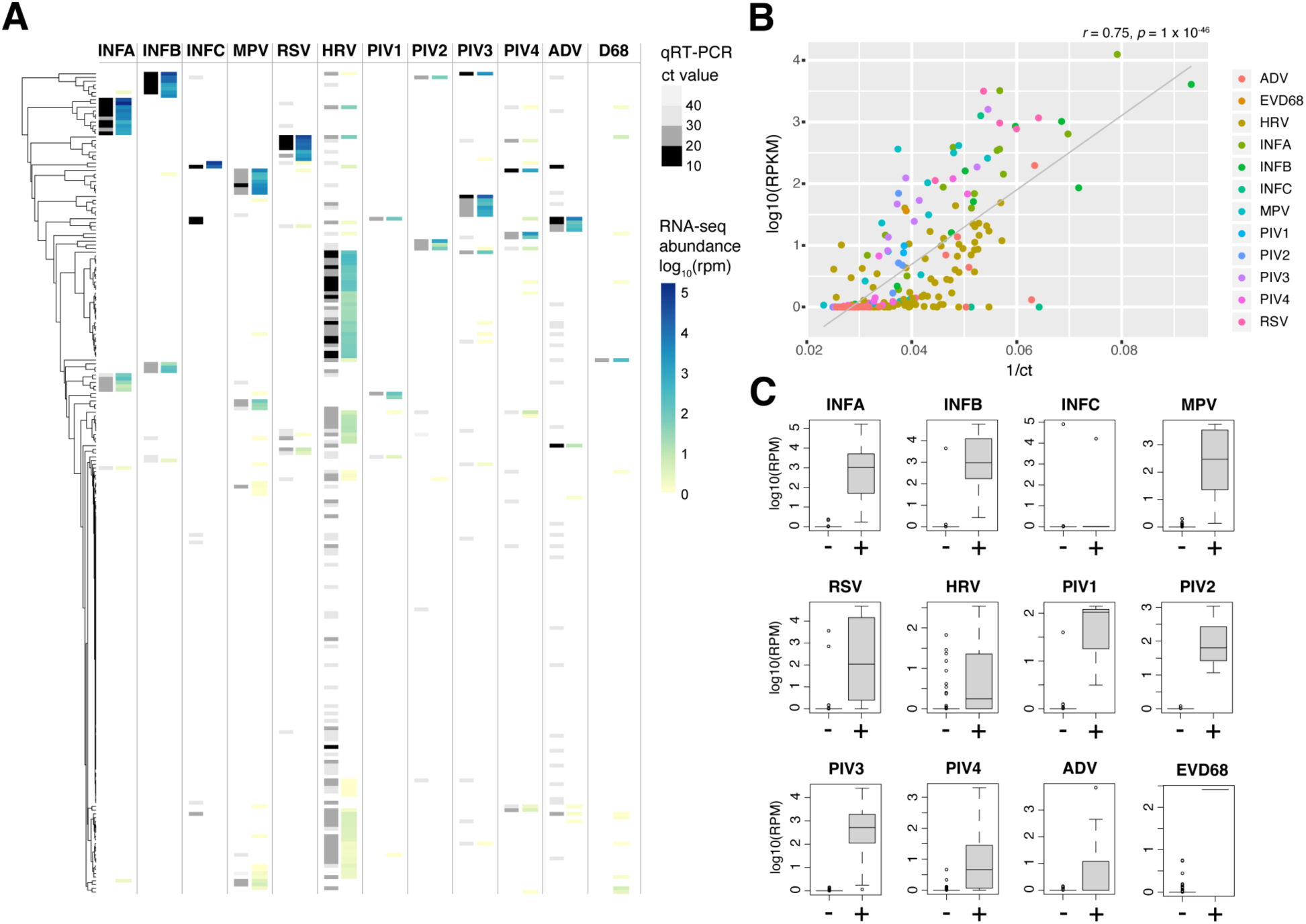
Detection of common respiratory viruses in NP metatranscriptomes. (**A**) Abundance heatmap for viruses detected in NP metatranscriptomes for 221 patients. For each virus, the qRT-PCR result is shown on the left of the column as a color gradient (negative – white, high to low cycle threshold values – light gray to black), and the estimated RNA-seq abundance is depicted on the right of the column as a color gradient (absent – white, low – yellow, high – dark blue). Each row in the heatmap and tip in the hierarchical tree corresponds to an individual patient sample. (**B**) qRT-PCR abundance (1/ cycle threshold) versus metatranscriptomic viral load (log_10_ of the RPKM). The estimated viral load from RNA-seq is significantly correlated with 1/Ct value from qRT-PCR. (**C**) Metatranscriptomic abundance of respiratory viruses in negative (−) versus positive (+) samples (labelled on X axis) defined by qRT-PCR test result. The boxes show the interquartile range and median line, and the whiskers show the variability extending to the furthest data points within 1.5 times above and below the interquartile range. Outliers outside of these ranges are shown as data points.

We then calculated the accuracy of viral detection by using the results of the qRT-PCR tests as the ground truth. Due to the uniqueness of viral sequences, we found that a very low threshold (>=1 RPM) was sufficient to distinguish virus-positive from negative samples. Using this threshold, we calculated the sensitivity and specificity of metatranscriptomic pathogen detection for each of the 12 viruses as shown in Table 2. Nine out of the 12 viruses were detected with 90-100% sensitivity and specificity, while INFC, HRV, and ADV were detected with lower accuracy. Overall, we were able to detect the 12 viruses with an average sensitivity/specificity of 86%/92%. These accuracies are consistent with other studies performing sequencing-based pathogen detection using NP samples (*27, 28*).

### RNA-seq uncovers additional pathogens and alternate explanations of disease etiology

By sequencing total RNA within a sample, metatranscriptomics has the potential to detect additional pathogens beyond those tested by culture or qRT-PCR. We therefore screened our RNA-seq dataset for additional pathogens previously associated with URTIs and/or sinusitis infections, as well as non-URTI pathogens and opportunistic pathogens, and further validated the identified species using additional bioinformatic approaches (see Methods). Across the 221 patient samples, we detected 22 additional pathogens that were not tested for clinically, including 11 bacteria and 11 viruses (Fig. 4). These species were then ranked in terms of their maximum relative abundance within a sample (Fig. 4).

**Fig. 4.**
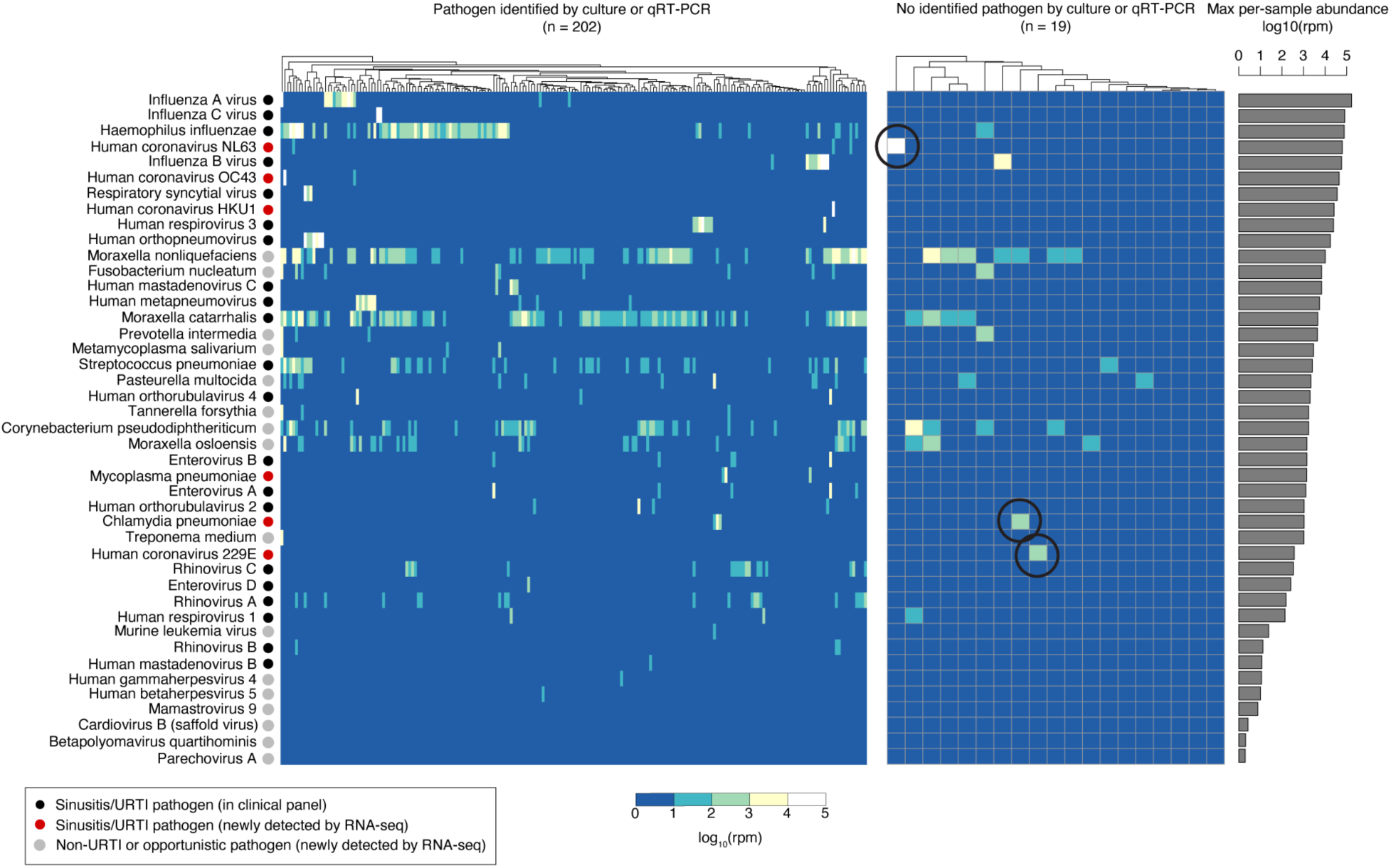
Metatranscriptomics of NP samples from children with acute sinusitis identified organisms not detected by qRT-PCR or culture. The organisms included in the heatmap are a subset of the full set of organisms detected by RNA-seq that exceed minimum abundance thresholds and include human pathogenic bacteria and viruses (see table S6 for full dataset). The organisms are sorted vertically based on their maximum relative abundance within a sample (across 221 samples). The heatmap displays the relative abundance of each organism in each sample as estimated by Kraken 2. The left heatmap includes samples with clinically identified pathogens by qRT-PCR or culture (N = 202), and the right heatmap includes 19 samples without a pathogen detected by qRT-PCR or culture. For the latter samples, several samples contain additional organisms identified by metatranscriptomics that are plausible causes of sinusitis. The barplot on the right depicts the total number of samples containing each detected pathogen.

Newly identified bacterial pathogens includes fourteen species listed in Fig. 4. The most notable identifications include *Mycobacterium pneumoniae* and *Chlamydia pneumoniae*, which were not included in the clinical panel but have been previously implicated in pediatric sinusitis and URTIs (*29, 30*). In addition, opportunistic pathogens including *Fusobacterium nucleatum*, *Moraxella* spp., and others, were also detected (Fig. 4), but some of these likely have a commensal role in the nasopharynx. Interestingly, we also detected periodontitis-associated bacteria, *Treponema medium*, *Prevotella intermedia*, and *Tannerella forsythia* (*31*), in a few (N = 1 to 4) samples, and all three co-occurring in the same patient. Follow-up investigation of this patient revealed that they were admitted to an emergency room with a severe tooth infection one year after the NP swab sample was taken, highlighting the potential of NP RNA-seq to detect subclinical infection.

Newly identified viral pathogens with the highest abundance include four human coronaviruses known to cause upper respiratory infections (NL63, OC43, HKU1, and 229E). We also detected parechovirus A and cardiovirus B (saffold virus), which have been associated with respiratory illness in children (*32, 33*), as well as other viruses that are not typically associated with respiratory infections including mamastrovirus 9, enteroviruses A and B, human gammaherpes virus 5, human betaherpes virus 5, and sequences related to murine leukemia virus (Fig. 4).

Of the 19 samples that had no pathogen detected by culture or qRT-PCR, 11 contained identified pathogens based on RNA-seq profiling. Three of the 11 samples (circled in Fig. 4) contained known pathogens detected at high abundance that were not included in the clinical pathogen panel: the coronaviruses NL63 and 229E, and the bacterium, *Chlamydia pneumoniae*. Eight of the 11 samples had pathogens detected by RNA-seq but not by qRT-PCR or culture, including influenza B (N = 1), parainfluenza virus 1 (N = 1), SPN (N = 1), MCAT (N = 4), and HFLU (N = 1).

Ultimately, these additional detected pathogens highlight the ability of RNA-seq to provide a more complete picture of the microbiome and virome present in acute sinusitis samples and suggests an expanded panel of viruses and bacterial pathogens to be used in future clinical workflows.

### Viral genome assembly and subtyping from host-derived metatranscriptomes

Read-based taxonomic classifications provide an estimate of microbial species present in each sample. However, *de novo* genome assembly methods may be used to assemble longer fragments including genomes of full-length RNA viruses, which can validate read-based predictions and reveal additional information.

By aligning the RNA-seq reads to reference genomes of identified viruses, we were able to assemble partial to complete genomes for a total of 205 viruses across 163 samples, including 25 different human pathogenic viruses (Fig. 5A). In addition to the 12 viral groups from the clinical panel (Fig. 3), genomes were assembled for 9 additional respiratory viruses (e.g., coronaviruses) not tested for clinically. We also assembled genomes of enterovirus A and B, WU polyomavirus, and mamastrovirus 9, which are typically implicated in other illnesses such as gastroenteritis. A total of 31 (15%) were 100% complete, while 60 (30%) had completeness >90% (table S7). All assembled viral genomes were phylogenetically verified by sequence comparison to related genomes in NCBI through BLAST, with average nucleotide identities (ANIs) ranging from 95-100%.

**Fig. 5.**
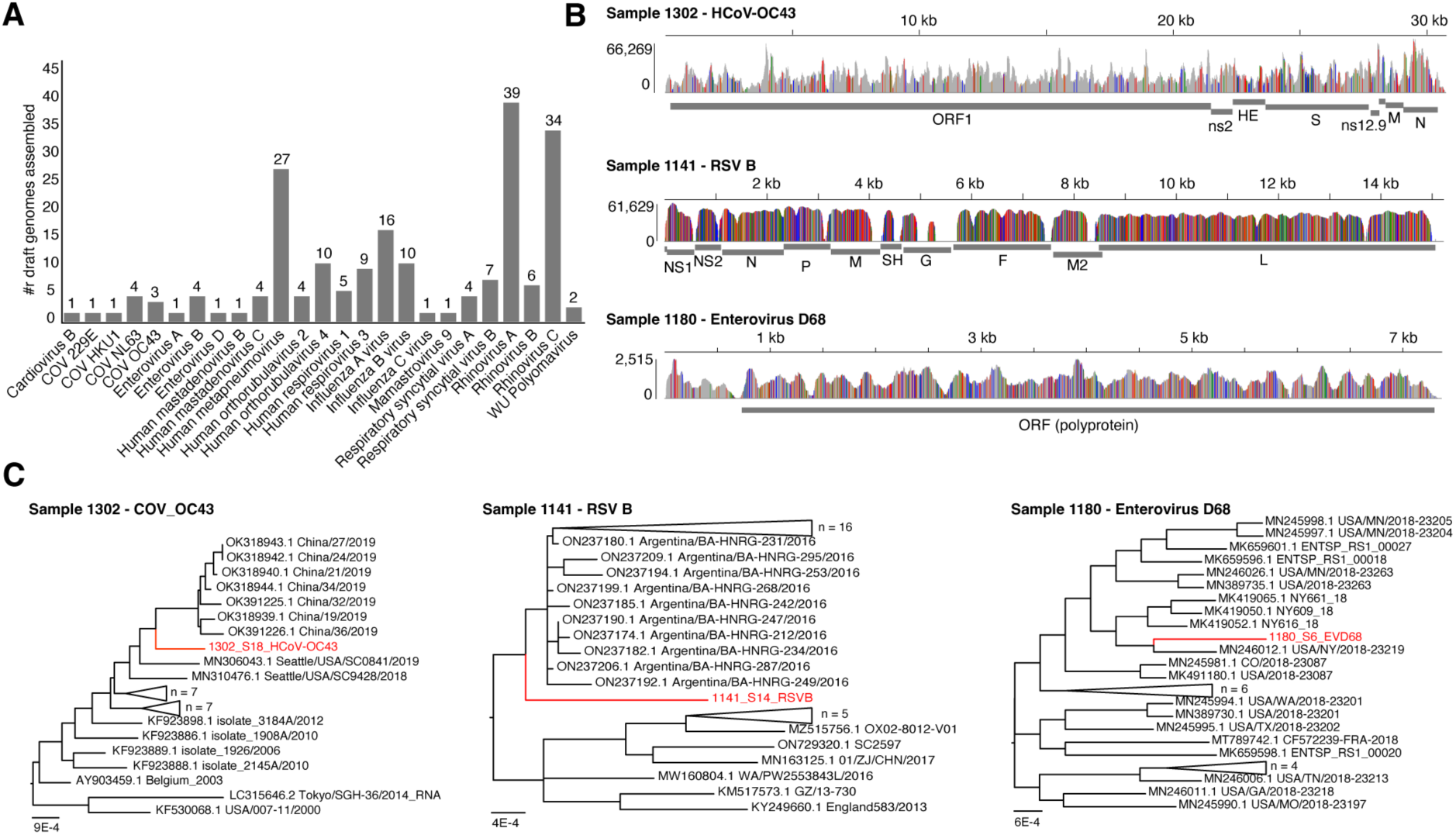
Assembled genomes of viruses from children with clinically diagnosed acute sinusitis. (**A**) Bar graph depicting the number of assembled genomes for various species of respiratory viruses across the full dataset (N = 205 total viruses assembled from 163 samples). (**B**) Read pileups for three selected samples showing sequencing reads mapped to reference genomes of human coronavirus (HCoV) OC43, RSV, and enterovirus D68 with SNP profiles as colored lines. (**C**) Phylogenetic analysis of three assembled viral genomes and their top 25 closest matching complete genomes from BLAST. Each newly assembled virus (red) is a unique strain that clusters as a distinct branch within its phylogenetic tree.

To explore the use of assembled genomes for viral subtyping, we focused on the predictions for influenza A and B, since these were subtyped clinically using qRT-PCR. The subtyping results using assembled influenza genomes showed excellent agreement with the clinical results, with Influenza A subtypes H1N1 and H3N2 having 100% (15/15) agreement and Influenza B subtypes Yamagata and Victoria having 82% agreement (9/11) with qRT-PCR results (table S8).

We then focused on several cases of interest, performing a deeper genomic and phylogenetic analysis of newly assembled genomes. Three examples of assembled viral genomes are shown in Fig. 5B, including a genome of a novel HCoV-OC43 strain, an RSVB genome, and an enterovirus D68 genome. All three of these genomes have distinct mutation profiles from other strains in the NCBI database (Fig. 5B), and clustered as unique strains in phylogenetic analysis (Fig. 5C). All three of the genomes also showed broad sequencing coverage across the genome, with the exception of the RSVB genome from sample 1141, which showed a lack of coverage spanning the glycoprotein G gene. Interestingly, a previous study also identified G protein deletion mutant RSV strains in pediatric pneumonia patients from South Africa (*34*).

### Host-response expression profiles distinguish bacterial from viral infections

Although RNA-seq analysis was capable of detecting pathogens directly from reads, most reads within RNA-seq samples were host (human) derived, ranging from 64.7-99.9%, which enables host-response profiling to potentially identify host biomarkers and immune responses associated with disease etiology (*35–38*).

To identify differentially expressed genes (DEGs) associated with bacterial versus viral infections, we compared host gene expression profiles of patients with bacterial pathogens to those with viral pathogens based on clinical diagnostic testing (Fig. 6A). Due to the presence of many (N = 138) complex samples containing a mixture of viral and bacterial pathogens, we chose to simplify the initial comparison and compared samples with only bacterial pathogens (N = 33) to those with only viral pathogens (N = 31), but subsequently analyzed all 221 samples. A total of 821 significant DEGs were detected with *q* < 0.001, of which 548 genes had increased expression in bacterial-positive patients and 273 genes had increased expression in viral-positive patients (Fig. 6A, table S9). We termed these genes as “bacterial upDEGs” and “viral upDEGs”.

**Fig. 6.**
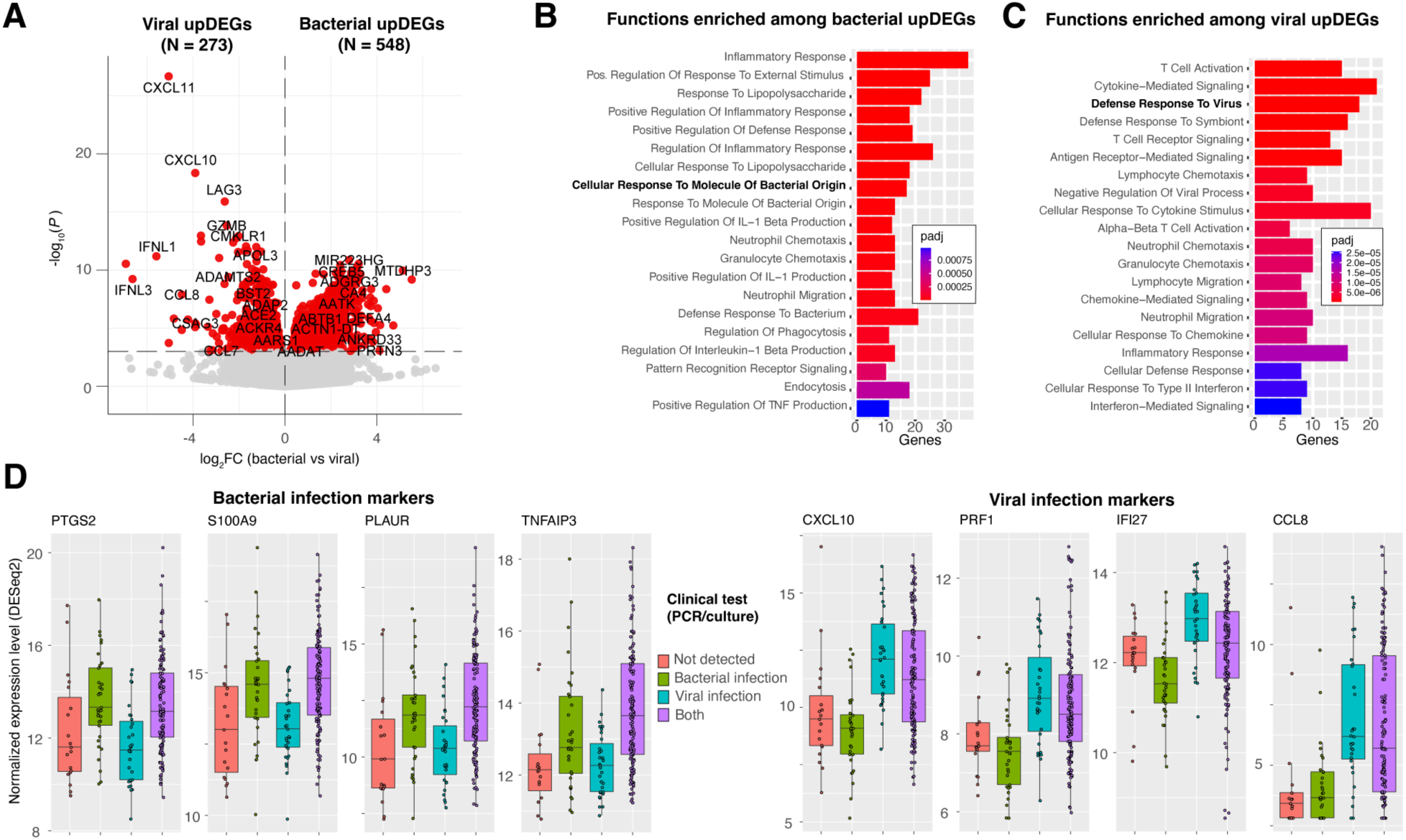
Identification of differentially expressed host genes indicative of host-responses to bacterial and viral infection in acute sinusitis patients. (**A**) Volcano plot of differentially expressed genes between samples with only bacterial pathogens and samples with only viral pathogens according to qRT-PCR and culture test results. Human genes shown in the upper right quadrant have significantly increased transcript abundance in samples with bacteria (bacterial upDEGs), and genes in the upper left quadrant have significantly increased transcript abundance in samples with virus(es). Genes are partitioned in the plot based on *p*-value significance thresholds. (**B** and **C**) Biological functions and pathways that are significantly enriched among bacterial and viral upDEGs, calculated using enrichR. For each function term, the associated adjusted p-value and number of genes is depicted. (**D**) Example bacterial and viral upDEGs and their expression levels (transcript abundance) across four categories of patients: those with neither bacteria or virus detected by culture or qRT-PCR; those with only bacteria, those with only virus, and those with both a bacteria and virus.

Based on function enrichment analysis, bacterial upDEGs were significantly associated with neutrophil regulation, regulation of inflammatory response, response to lipopolysaccharide, and response to molecule of bacterial origin (Fig. 6B), which are consistent with an immune response to bacterial infection. The identified bacterial upDEGs include genes previously shown to be markers of bacterial infection: for example, PTGS2 (6-fold increase in bacterial-positive patients, *q* = 3.1 x 10^-7^), S100A9 (4-fold increase, *q* = 4.2 x 10^-6^, PLAUR (5-fold increase, *q* = 7.3 x 10^-6^), TNFAIP3 (4-fold increase, *q* = 1.3 x 10^-5^), IL1A (6-fold increase, *q* = 1.0 x 10^-4^), IL1B (6-fold increase, *q* = 4.0 x 10^-5^), CXCL2 (4-fold increase, *q* = 1.3 x 10^-5^), and NFKBIA (4-fold increase, *q* = 1.8 x 10^-5^) (Fig. 6D).

Viral upDEGs were found to be significantly associated with cytokine signaling, defense response to virus, T cell receptor signaling, and inflammatory response (Fig. 6C), which are related to viral immune response pathways. Consistent with this, the identified viral upDEGs include genes shown to be markers of viral infection in previous studies: for example, CXCL11 which was increased 33-fold in virus-positive patients (*q* = 4.9 x 10^-23^), CXCL10 (15-fold increase, *q* = 2.6 x 10^-15^), CCL8 (23-fold increase, *q* = 2.3 x 10^-6^), PRF1 (4-fold increase, *q* = 3.8 x 10^-9^) and IFI27 (2-fold increase, *q* = 8.5 x 10^-7^), which represent putative biomarkers of viral infection in our analysis (Fig. 6D).

In general, representative viral and bacterial upDEGs had lower expression levels for samples in which no bacteria or virus was detected by qRT-PCR/culture, and higher expression levels for samples containing both a virus and bacterial pathogen (Fig. 6D). Interestingly, there are several exceptions to this pattern including four samples that had a strong antiviral response despite there being no virus detected by qRT-PCR/culture. Deeper investigation of these samples by RNA-seq revealed that three of them contained respiratory viruses (two coronaviruses and influenza B) (Fig. 4B) that were not detected by the qRT-PCR tests. Other exceptions include two samples which had no bacterial pathogen detected by culture/qRT-PCR but had a strong antibacterial response. One of these samples (sample 1303) had a bacterial pathogen (MCAT) identified in high abundance by RNA-seq. These results suggest that host-response profiling may provide an indication of viral or bacterial infection when traditional tests fail to detect a pathogen.

### Magnitude of host responses correlates with viral and bacterial pathogen abundance

If the identified viral and bacterial upDEGs are genuine biomarkers of viral and bacterial infections, respectively, then their levels of expression should correlate with the abundance of viral and bacterial pathogens estimated from RNA-seq. To test this hypothesis, we calculated the total bacterial pathogen abundance as the sum of the relative abundance of the pathogens SPN, HFLU, and MCAT. We then binned all samples into ten groups, with group 1 having the lowest bacterial pathogen abundance, and group 10 having the highest. We then repeated this analysis for viral pathogens, summing the total abundance of 12 viral pathogens as well as the coronaviruses that were clearly present based on RNA-seq data, but missing from the clinical test.

As shown in Fig. 7A, with increasing abundance of bacterial sinusitis pathogens (MCAT, SPN, HFLU), there is a clear increase in expression levels of bacterial upDEGs. To quantify this pattern, for each sample we calculated the “magnitude” of the bacterial and viral host response as the average expression level (Z-score) of the bacterial and viral upDEGs. As shown in Fig. 7B, the magnitude of bacterial host response correlated significantly with bacterial pathogen abundance (Pearson *r* = 0.50, two-tailed *p* = 1.6 x 10^-15^). The same pattern was also seen for viruses: that is, the abundance of viral pathogens also correlated significantly with the magnitude of viral host-response (Pearson *r* = 0.33, two-tailed *p* = 5.8 x 10^-7^) (Fig. 7C,D). Both the bacterial and viral host responses however did not correlate with other clinical features including the duration of cold symptoms and symptom severity (Fig. 7A). Although these pathogen-host-response correlations are a general pattern, not all samples display this trend. For example, several samples with high bacterial pathogen abundance lack a strong bacterial host response. In addition, one outlier (marked * in Fig. 7A) shows an individual with a low detected bacterial pathogen abundance but a strong bacterial host response. This could indicate an immune response to an unknown bacterial species.

**Fig. 7.**
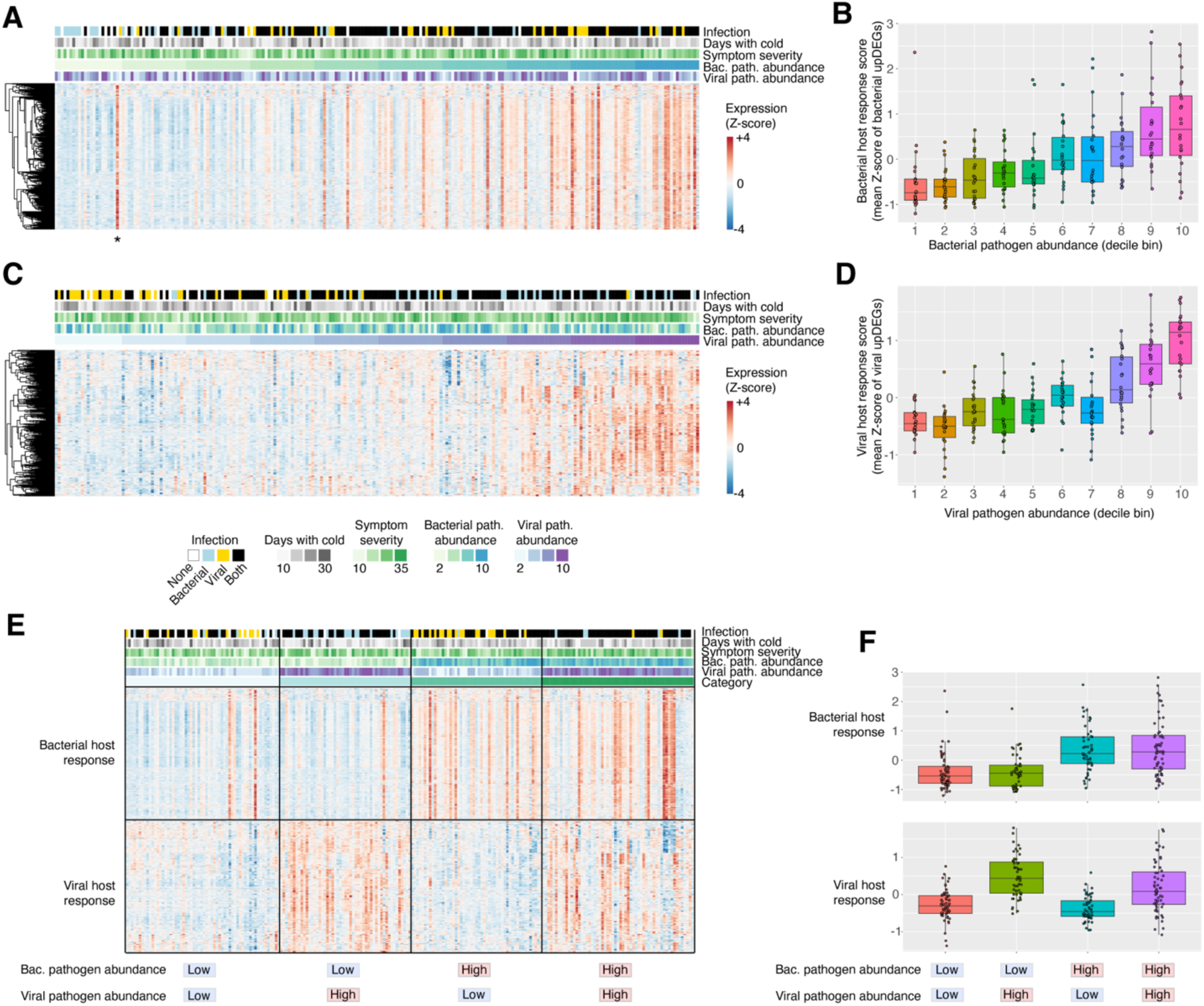
Host-response correlates with relative abundance of bacterial and viral pathogens. (**A**) Expression heatmap of bacterial upDEGs (bacterial host response genes), with samples (columns) sorted by total metatranscriptomic bacterial pathogen abundance. The associated metadata for all samples is also plotted above the heatmap. * Also shown is an outlier sample associated with a strong bacterial host response but with low detected abundance of MCAT, HFLU, or SPN. (**B**) Bacterial host response score versus metatranscriptomic bacterial pathogen abundance. The bacterial host response score was calculated as the mean expression level (Z-scores) of all the bacterial upDEG genes. (**C**) Expression heatmap of viral upDEGs (viral host response genes), with samples (columns) sorted by metatranscriptomic viral pathogen abundance. (**D**) Viral host response score versus metatranscriptomic viral pathogen abundance. The viral host response score was calculated as the mean expression level (Z-scores) of all the viral upDEG genes. (**E**) Heatmap of bacterial and viral host responses (upDEGs), where samples (columns) have been sorted into four groups based on high or low bacterial/viral pathogen abundance, with high considered as a 60^th^ percentile or greater relative abundance. In general, samples with low bacterial and viral abundance tend to lack a bacterial/viral host response, whereas samples containing bacteria, viruses, or both displayed the appropriate response. (**F**) Jitter plots of the bacterial and viral host response scores across four categories of samples. Bacterial and viral host response scores were calculated by averaging the expression level Z-scores of all bacterial and viral upDEGs, respectively.

In addition to the association between host-response and pathogen abundance, we also tested for host-response correlations with other clinical metadata. A weaker but significant (*r* = 0.33, *p* = 6.6 x 10^-7^) host-response pattern was detected between a subset of genes and patient symptom severity scores (PRSS) at the time of diagnosis. A total of 45 genes were differentially expressed as a function of PRSS, which subdivided into 2 expression clusters (fig. S2). Cluster 1 was positively correlated with PRSS and includes the following genes: *METTL7B, MMP3, PRF1, GNLY, MMP1, FPR3, GIMAP6, OLFML2B, DESI1, IL12RB2*. Function enrichment analysis revealed that cluster 1 was associated with a response to infection (cellular defense response, natural killer cell mediated immunity, and cellular response to cytokine stimulus). Other pathways such as proteolysis and pyroptosis are also involved in innate host immune response by eliminating and degrading infected cells (*39, 40*).

### RNA-seq classifies patients into distinct groups with unique pathogen-host response profiles

After examining host responses to bacterial and viral infections individually, we considered how bacterial and viral relative abundance together impact host responses within patients. To investigate this, we used the RNA-seq abundance to bin samples into four groups: those with *low bacterial / low viral* pathogen abundance (N = 60, 27%), *high viral / low bacterial* pathogen abundance (N = 51, 23%), *high bacterial / low viral* pathogen abundance (N = 51, 23%), and *high bacterial / high viral pathogen* abundance (N = 59, 27%). Here, the thresholds of “high” and “low” pathogen abundance based on RNA-seq estimated levels (>=60^th^ percentile) and not the presence/absence classification obtained from qRT-PCR and culture-based testing.

The four groups of patients display distinct host response signatures (Fig. 7E,F). As expected, samples with low bacterial and low viral pathogen abundance tend to have weak bacterial and antiviral responses (Fig. 7E). Samples with high viral abundance but low bacterial abundance display a strong antiviral pattern and a weak bacterial response. Samples with high bacterial pathogen abundance but low viral pathogen abundance are associated with a strong bacterial host response, and samples with high bacterial and viral pathogen abundance show both host responses. Again, there are several outliers that are exception to these general trends. The viral host response for individuals with both bacterial and viral pathogens was lower than the viral-only group (*p* = 0.01), and the bacterial host response for individuals with both bacterial and viral pathogens was not significantly different from the bacterial-only group (*p* = 0.82). Finally, we tested whether the bacterial and viral host-response magnitude alone could predict samples with high pathogen abundance, with pathogen abundance defined as described above using RNA-seq measurements. The bacterial host response magnitude predicted high-bacterial samples with an AUROC of 0.79, and the viral host response magnitude predicted high-virus samples with an AUROC of 0.80. If sensitivity is desired over specificity, high-bacterial samples could be predicted with a sensitivity/specificity of 80%/68% using host-response information alone. Ultimately, these analyses suggest that host-response information alone may have diagnostic value in differentiating between viral and bacterial sinus infections, especially when the relative abundance (pathogen load) is high.

## DISCUSSION

In this study, we performed metatranscriptomic analysis of 221 NP samples from children with clinically diagnosed acute sinusitis. Prior to this work, there has been a lack of research evaluating the use and applications of RNA-seq profiling in this clinical context. Our study provides several research contributions. First, it highlights the ability of RNA-sequencing of clinical samples to accurately identify bacterial and viral pathogens associated with sinusitis infections and URTIs. Second, it provides an original dataset to assist with the development of future bioinformatic approaches for infectious disease profiling, including hundreds of assembled viral pathogen genomes contributing to ongoing pathogen genomic surveillance efforts. Third, it identifies host-response signatures of bacterial and viral infections in sinusitis, which could serve as the basis for the development of biomarker assays to be used in future clinical workflows that optimize delivery of care.

Using RNA-seq we achieved an overall sensitivity of 87% and specificity of 81% in reproducing the clinical results for detection of three bacterial species that are mostly commonly implicated in sinusitis (*4*). RNA-seq also demonstrated a significant ability to detect viral pathogens that were also detected by the qRT-PCR panel (average sens/spec of 86%/92%), as well as predict viral load (Ct value). These accuracies are comparable to results obtained by previous studies using NGS for pathogen detection in NP samples (*27, 28*).

For clinical decision making regarding antibiotic treatment, a key goal of sequencing-based approaches is to not only detect the pathogen of interest but also its antimicrobial genes, which can be especially challenging in mixed metagenomic samples. As proof of principle, we focused on beta-lactamase resistance in HFLU isolates, which represents a key clinical issue (*41, 42*). As done previously for pediatric nose and ear samples (*43*) we used CARD (*25*) to identify beta-lactamases in RNA-seq data. This RNA-seq workflow was able to correctly detect beta-lactamase genes in 67% of the resistant HFLU isolates, with a specificity of 96%. Additionally, beta-lactam resistance SNPs in the *Haemophilus influenzae* PBP3 gene were also detected in several samples, which may represent an additional resistance mechanism that was detected by RNA-seq profiling but not covered by clinical AMR testing.

Finally, through *de novo* assembly methods, we were able to assemble genomes of 205 viral pathogens with varying degrees of completeness. Assembled genomes confirm read-based predictions and provide added information that cannot be obtained from short sequencing reads or qRT-PCR-based methods. For example, phylogenetic analyses of some of these viruses (e.g., HCoV-OC43, RSV B, enterovirus D68) revealed unique differences from closely related genomes in the database, suggesting that they represent distinct strains. A potentially relevant mutation (absence of large segments of the G gene) was identified in an RSV B strain similar to previous reports (*34*). Further analysis of RSV genomes from patient samples is needed to determine the frequency of G deletion mutants, which could be important information to consider for RSV vaccine design.

An advantage of metatranscriptomic RNA-seq over culture or qRT-PCR is the ability to perform a broad and untargeted analysis to detect any species whose genome is available in the reference database, which theoretically improves sensitivity of pathogen detection and discovery. Out of 221 pediatric sinusitis patients tested, 19 did not have any bacterial or viral pathogen detected by culture-based or qRT-PCR testing. RNA-seq identified plausible pathogens for acute sinusitis in 11 of these 19 samples including cases of influenza B and PIV1 that were missed by qRT-PCR. Not surprisingly, several new pathogenic bacteria and viruses were also detected in these samples and were verified by genome assembly and phylogenetics. These included two coronaviruses (NL63 and 229E), as well as the bacterium, *Chlamydia pneumoniae*. Other identified organisms included commensal organisms of the nasal microbiome and opportunistic pathogens that may or may not play a direct role in sinusitis (e.g., different species of *Moraxella* and *Corynebacterium*). Clarifying the role of these and other species in sinusitis etiology is a challenging goal for future work.

One of the most exciting aspects of this study is the identified host-response gene expression patterns associated with bacterial and viral sinusitis infectious. Since the pathogen composition of our patient cohort was complex including a large number of samples containing both bacterial and viral pathogens based on culture/qRT-PCR, we chose to simplify the initial comparison between virus-positive only samples versus bacteria-positive only samples. This enabled the detection of virus associated and bacteria associated host DEGs (“viral host response” and “bacterial host response”) that formed the basis of subsequent analyses. Remarkably, the magnitude of these host responses correlated significantly with the total abundance of bacterial or viral pathogens detected in the samples. Importantly, this correlation between pathogen abundance and host-response magnitude was only identified for a limited subset of bacterial species (those previously identified as sinusitis pathogens, MCAT, SPN, HFLU) and respiratory viruses, and the correlation was absent when examining other species detected in the data that may reflect commensal organisms. This finding indicates that the relative abundance of specific bacterial and viral species within the nasopharynx is a determinant of the strength of the host immune response. This is consistent with immunology since the expression of host antiviral and antibacterial pathways are dependent on the levels of viral (e.g., dsRNA) and bacterial pathogen-associated molecular patterns (e.g., lipopolysaccharide) sensed by the host immune system. Previous studies have also reported a correlation between antiviral host responses in RNA-seq and viral load (*44–46*). However, our study is unique by analyzing the interplay between a complex mixture of bacterial and viral pathogens and their impact on the host transcriptomic response.

Although traditional methods (culture and qRT-PCR) provided a simple classification of our samples based on detected presence/absence of a pre-defined set of pathogens, metatranscriptomic data enabled a more holistic classification based on pathogen abundance and host-response information (Fig. 7). When taking both pathogen abundance and host-response information into consideration, the samples could be subdivided into four main groups: those with a “low” abundance of bacterial or viral pathogens which tend to lack a host-response, and those with a “high” abundance of bacterial pathogens, respiratory viruses, or both, which tend to show the expected host responses. Interestingly, the observed correlation between pathogen abundance and host-response is not perfect; there are several outlier samples which exhibited a strong host-response pattern and yet lack a detected pathogen, and other samples which contained a high pathogen abundance but lack a detectable host response. For the former category, it is possible that those samples contained other pathogens that were not included in our pathogen panel, which may include opportunistic infections by commensal organisms for example. For the latter category, these cases could indicate delayed host-responses in patients at the time of sampling, shedding of viral RNA at a post-infection time point which may be associated with a reduced host-response, or simply an imperfect correlation between host-responses and pathogen abundance. Nevertheless, future research focusing on host-responses of patients with infectious disease and factors that account for discrepancies between detected pathogen abundance could clarify mechanistic understanding of disease etiology.

There are several limitations of our study that could account for variation in the results obtained. First, the classification into viral and bacterial infection was inferred based on the presence/absence of bacterial and viral pathogens, but some of these organisms may be present as commensals and their presence alone does not necessitate an infection (*47, 48*). Second, the enrollment criteria for this study recruited patients experiencing symptoms for at least 6 days when sampled. Since peak shedding of some viruses can occur within 48 hours of symptom onset, the chosen sampling time may have led to a reduced sensitivity of viral detection as well as lower coverage for genomes assembled. Variation in the timing of bacterial infections could also impact sensitivity of bacterial detection by RNA-seq. Third, our sensitivity for pathogen detection by RNA-seq is dependent on the depth of sequencing. Deeper sequencing may have been necessary to detect viruses, for example, that were false negatives by RNA-seq but were detected using qRT-PCR. DNA viruses in particular (e.g., adenoviruses) may have been more prone to weak detection due to the use of RNA-seq over DNA-seq. Future studies that employ both metatranscriptomic and metagenomic sequencing with repeated time-series sampling of patients may overcome some of the limitations described above. Nevertheless, the current study provides a starting framework for exploring the use of high-throughput sequencing of patient samples to uncover etiology and host-response in pediatric sinusitis and other upper respiratory infections.

## MATERIALS AND METHODS

### Study design and description of the cohort

Between February 2016 and April 2022, 510 children 2 to 11 years of age (inclusive) with clinically diagnosed acute sinusitis were enrolled in a randomized multicenter double-blind trial (ClinicalTrials.gov number, NCT02554383). Exclusion criteria have been previously described (*6*). Children were recruited from 6 outpatient centers. Children were randomly assigned to receive 10 days of amoxicillin-clavulanate or matching placebo. A total of 204 patients did not have a NP sample collected, or their sample was not preserved in RNA buffer and were excluded. Of the remaining 306 patients’ samples, 61 were not sequenced due to low RNA yield. Although 245 samples underwent RNA-sequencing, batch 1 was prepared with a different kit/protocol and when analyzed displayed a strong batch effect and was thus removed, leaving 221 patients. The primary outcome, symptom burden, was assessed by having parents complete the Pediatric Rhinosinusitis Symptom Scale (PRSS) electronically every evening on Days 2 to 11. As previously described (*6*) the PRSS is a validated scale that assesses symptoms of sinusitis.

### Culture and sensitivity pattern of bacterial pathogens

We collected NP swabs from all children at study entry. As previously described (*23*), the tip of the swab was cut, placed in DNA/RNA shield (Zymo, R1100), and transported on ice to the lab. The remainder of the swab was placed into Amies transport medium and transported on ice to the Clinical Laboratory at UPMC Children’s Hospital of Pittsburgh within 48 hours and plated on blood and chocolate agars. Identification of SPN, HFLU, and MCAT on culture was accomplished using standard microbiological techniques. HFLU isolates were tested for the beta-lactamase production using a cefinase disk.

### qRT-PCR for viral co-infection

Using an aliquot of Amies transport media plus MagMax lysis/binding buffer, nucleic acid extraction was performed for viral identification using the ABI MagMax96 Express automated instrument and the MagMax 96 Viral Isolation Kit (Thermo Fisher, AMB 18365) (*23, 49*). Adenovirus, influenza subtypes A/B/C, human metapneumovirus (HMPV), human rhinovirus (HRV), parainfluenza virus (PIV) subtypes 1-4, Enterovirus D68, and respiratory syncytial virus (RSV) were tested for using individual real-time qRT-PCR assays. A Ct threshold of 40 was used for all viruses and positive and negative controls were included in each run.

### RNA-seq library generation, sequencing, and data processing

RNA was assessed for quality using a Fragment Analyzer 5300 and RNA concentration was quantified on a Qubit FLEX fluorometer. Libraries were generated with either the Illumina TruSeq Stranded Total RNA prep (20020599) or the Illumina Stranded Total Library Prep kit (Illumina: 20040529) according to the manufacturer’s instructions, after using the Illumina Ribo-Zero Plus rRNA Depletion Kit (20037135). Batch 5 was additionally treated with the Illumina Ribo-Zero Plus Microbiome rRNA Depletion Kit (20072062). For library generation, 100 ng of input was used for the Illumina TruSeq Stranded Total RNA protocol with 15 cycles of indexing PCR, and 20-100 ng of RNA input was used for the Illumina Stranded Total Library Prep protocol with 15 cycles of indexing PCR for 100ng of RNA input and 17 cycles of indexing PCR for input RNA ≤100 ng. Library quantification and assessment was done using a Qubit FLEX fluorometer and the Fragment Analyzer 5300. Libraries were normalized and pooled to 2 nM by calculating the concentration based off the fragment size (base pairs) and the concentration (ng/μl) of the libraries. Sequencing was performed on an Illumina NextSeq 2000, using a P3 200 flow cell with sequencing read lengths of 2×101bp, with a target of 40 million reads per sample. Sequencing data was demultiplexed by the Illumina on-board DRAGEN FASTQ Generation software. Library generation and sequencing was performed by the University of Pittsburgh Health Sciences Sequencing Core (HSSC), Rangos Research Center, UPMC Children’s Hospital of Pittsburgh, Pittsburgh, Pennsylvania, United States of America.

Fastp v0.23.1 (*50*) was used for quality trimming and adapter removal on default parameters. FastQC v0.11.9 (*49*) and MultiQC v1.12 (*51*) were used to check the quality of all sequence files before and after processing to ensure data was ready for analysis.

### Taxonomic classification of RNA-seq reads for detection of bacterial and viral pathogens

Taxonomic classification of sequencing reads was performed using Kraken 2 v2.1.2 (*24*) with default parameters. The PlusPF database dated 9/8/2022 (https://benlangmead.github.io/aws-indexes/k2) was used with Kraken 2, which was originally built from NCBI RefSeq archaeal, bacterial, viral, plasmid, human, UniVec_Core, protozoan, and fungal sequences. A Kraken 2 detection threshold of 3 reads was used for bacterial species (selected based on F1 score optimization), while no threshold was used for viruses. New pathogens identified by Kraken 2 but not included in the clinical panel were further validated using BLAST (*52*), MASH (*53*) and metAnnotate (*54*), focusing on samples associated with the largest estimated abundance for each pathogen.

The normalized abundance of each taxon was calculated as the number of reads per million (RPMs). Relative abundance heatmaps were generated using R v4.2.1 and the pheatmap package. For display, log_10_(RPM + 1) values were used to avoid log(0) errors. Receiver operator curves were also generated in R and the area under the curve was computed using the pROC package. Pathogen abundance jitter plots and top species plots were generated using ggplot2 in R (*55*).

Viral load was estimated from RNA-seq data following the method of Graf et al (*27*). The number of detected reads for a virus was divided by the total number of reads in the sample and the size of the respective viral genome in kilobases, and then multiplied by 1 million to generate an RPKM value (reads per kilobase of reference sequence per million total sequencing reads).

### Detecting beta-lactamase genes using RNA-seq

For the samples that were positive for HFLU based on culture tests, sequencing reads classified as non-human by Kraken 2 were extracted using extract_kraken_reads.py and assembled into contigs using the rnaSPAdes v3.15.4 with default parameters (*56*). Using CARD resistance gene identifier (RGI) software v6.0.1 (*25*) and default database, the contigs were analyzed with the ‘main’ function of the RGI tool with the ‘low-quality’ and ‘include-nudge’ parameters. The results were filtered to keep “strict” or “perfect” hits to beta-lactamase genes, genes acting on antibiotics belonging to the penam drug class, and hits with at least 10.0% sequence coverage to the reference gene.

### Viral genome assembly and phylogenetic analysis

RefSeq genomes for all viruses of interest were downloaded from NCBI. Non-human reads were mapped to viral genomes using BBMap v38.86 (*57*) to create .bam files. The consensus sequence for each sorted mapping result was produced using samtools v1.16.1 with the ‘-a’ option. A python script was used to calculate whole genome coverage relative to the RefSeq viral genome. Genome coverage was considered complete if >= 99.5%. FastANI v1.32 was used to calculate the average nucleotide identity to the closest reference genome for each genome assembled.

Complete viral genomes were queried against the complete NCBI non-redundant nucleotide database using BLAST (*52*). Up to 35 top matching sequences were downloaded and aligned to the assembled genome using the MUSCLE algorithm (*58*). The multiple genome alignment was used to generate a phylogenetic tree with FastTree v2.1.10 (*59*), and FigTree v1.4.4 was used for tree visualization.

### Host response gene expression analysis

Host transcript abundance quantification was performed using Salmon v1.7.0 (*60*) with the Human Gencode v39 reference transcriptome. Differential gene expression analysis was performed using using DESeq2 and tximport in R (*61*). Related statistical analyses are described in the following section. Heatmaps were produced in R using pheatmap, v1.0.12 jitter plots using ggplot2 v3.3.6, and volcano plots using the EnhancedVolcano package v1.14.0.

### Statistical analysis

Differentially expressed genes (DEGs) were detected by comparing samples positive for viruses only versus samples positive for bacteria only based on culture or qRT-PCR testing. In the design formula for the ‘DESeqDataSetFromTximport’ function, we also controlled for potential confounding variables “batch number”, “sex”, and “age (scaled)”. Log2 fold changes and adjusted *p*-values (*q*-values) were calculated for all genes, and a significance threshold of *q* <= 0.05 was used to identify DEGs. Function enrichment analysis of genes with significantly increased expression in the viral and bacterial groups was performed using EnrichR (accessed June, 2023) (*62*) with the GO Biological Process 2021 ontology and an FDR threshold of 0.05.

### List of Supplementary Materials

Figs. S1 to S2

Table S1 to S9

## Data Availability

Code and processed data associated with this paper is available at https://github.com/doxeylab/Abumazen-et-al. Additional raw de-hosted sequencing data and gene expression data are available from the authors upon request.

## General

The authors would like to acknowledge the Digital Research Alliance of Canada for providing high-performance computing infrastructure that enabled bioinformatic data analyses presented in this manuscript.

## Funding

This work was supported by:

The National Institute of Allergy and Infectious Diseases grant U01AI118506 (NS).

The Natural Sciences and Engineering Research Council of Canada (NSERC): RGPIN-2019-04266 (ACD)

The Natural Sciences and Engineering Research Council of Canada (NSERC), Canada Graduate Scholarships – Master’s (NA)

Government of Ontario, Ontario Graduate Scholarship (NA)

Government of Ontario, Ministry of Colleges and Universities, COVID-19 Rapid Research Fund C-077-2423557 (JH, ACD)

## Author contributions

Conceptualization: ACD, JH, NS

Methodology: All authors

Investigation: NA, VC, BL

Visualization: NA, ACD

Funding acquisition: NS, ACD, JH, NA

Project administration: ACD, NS

Supervision: ACD, NS

Writing – original draft: ACD, NA

Writing – review & editing: All authors

## References

1. I. Brook, Acute sinusitis in children. Pediatr Clin North Am 60, 409–424 (2013).

2. A. K. C. Leung, K. L. Hon, W. C. W. Chu, Acute bacterial sinusitis in children: an updated review. Drugs Context 9 (2020), doi:10.7573/DIC.2020-9-3.

3. K. E. Fleming-Dutra, A. L. Hersh, D. J. Shapiro, M. Bartoces, E. A. Enns, T. M. File, J. A. Finkelstein, J. S. Gerber, D. Y. Hyun, J. A. Linder, R. Lynfield, D. J. Margolis, L. S. May, D. Merenstein, J. P. Metlay, J. G. Newland, J. F. Piccirillo, R. M. Roberts, G. V Sanchez, K. J. Suda, A. Thomas, T. M. Woo, R. M. Zetts, L. A. Hicks, Prevalence of Inappropriate Antibiotic Prescriptions Among US Ambulatory Care Visits, 2010-2011. JAMA 315, 1864–73 (2016).

4. E. R. Wald, G. J. Milmoe, A. Bowen, J. Ledesma-Medina, N. Salamon, C. D. Bluestone, Acute maxillary sinusitis in children. N Engl J Med 304, 749–54 (1981).

5. C. L. Charlton, E. Babady, C. C. Ginocchio, T. F. Hatchette, R. C. Jerris, Y. Li, M. Loeffelholz, Y. S. McCarter, M. B. Miller, S. Novak-Weekley, A. N. Schuetz, Y. W. Tang, R. Widen, S. J. Drews, Practical Guidance for Clinical Microbiology Laboratories: Viruses Causing Acute Respiratory Tract Infections. Clin Microbiol Rev 32 (2018), doi:10.1128/CMR.00042-18.

6. N. Shaikh, A. Hoberman, T. R. Shope, J.-H. Jeong, M. Kurs-Lasky, J. M. Martin, S. Bhatnagar, G. B. Muniz, S. L. Block, M. Andrasko, M. C. Lee, K. Rajakumar, E. R. Wald, Identifying Children Likely to Benefit From Antibiotics for Acute Sinusitis: A Randomized Clinical Trial. JAMA 330, 349–358 (2023).

7. D. M. Knipe, P. Howley, Fields Virology (Lippincott Williams & Wilkins, ed. 6, 2013).

8. I. Brook, Microbiology of sinusitis. Proc Am Thorac Soc 8, 90–100 (2011).

9. A. Tada, N. Hanada, Opportunistic respiratory pathogens in the oral cavity of the elderly. FEMS Immunol Med Microbiol 60, 1–17 (2010).

10. W. Gu, S. Miller, C. Y. Chiu, Clinical Metagenomic Next-Generation Sequencing for Pathogen Detection. Annu Rev Pathol 14, 319–338 (2019).

11. C. Y. Chiu, S. A. Miller, Clinical metagenomics. Nat Rev Genet 20, 341–355 (2019).

12. F. Xie, Z. Duan, W. Zeng, S. Xie, M. Xie, H. Fu, Q. Ye, T. Xu, L. Xie, Clinical metagenomics assessments improve diagnosis and outcomes in community-acquired pneumonia. BMC Infect Dis 21 (2021), doi:10.1186/S12879-021-06039-1.

13. R. Schlaberg, K. Queen, K. Simmon, K. Tardif, C. Stockmann, S. Flygare, B. Kennedy, K. Voelkerding, A. Bramley, J. Zhang, K. Eilbeck, M. Yandell, S. Jain, A. T. Pavia, S. Tong, K. Ampofo, Viral Pathogen Detection by Metagenomics and Pan-Viral Group Polymerase Chain Reaction in Children With Pneumonia Lacking Identifiable Etiology. J Infect Dis 215, 1407–1415 (2017).

14. L. Van Tan, N. Thi Thu Hong, N. My Ngoc, T. Tan Thanh, V. Thanh Lam, L. Anh Nguyet, L. Nguyen Truc Nhu, N. Thi Ha Ny, N. Ngoc Quang Minh, D. Nguyen Huy Man, V. Thi Ty Hang, P. Nguyen Quoc Khanh, T. Chanh Xuan, N. Thanh Phong, T. Nguyen Hoang Tu, T. Tinh Hien, L. Manh Hung, N. Thanh Truong, L. Min Yen, N. Thanh Dung, G. Thwaites, N. Van Vinh Chau, for OUCRU COVID-19 research group, SARS-CoV-2 and co-infections detection in nasopharyngeal throat swabs of COVID-19 patients by metagenomics. J Infect 81, e175–e177 (2020).

15. A. Piantadosi, S. S. Mukerji, S. Ye, M. J. Leone, L. M. Freimark, D. Park, G. Adams, J. Lemieux, S. Kanjilal, I. H. Solomon, A. A. Ahmed, R. Goldstein, V. Ganesh, B. Ostrem, K. C. Cummins, J. M. Thon, C. M. Kinsella, E. Rosenberg, M. P. Frosch, M. B. Goldberg, T. A. Cho, P. Sabeti, Enhanced Virus Detection and Metagenomic Sequencing in Patients with Meningitis and Encephalitis. mBio 12, e0114321 (2021).

16. A. Ramesh, S. Nakielny, J. Hsu, M. Kyohere, O. Byaruhanga, C. de Bourcy, R. Egger, B. Dimitrov, Y.-F. Juan, J. Sheu, J. Wang, K. Kalantar, C. Langelier, T. Ruel, A. Mpimbaza, M. R. Wilson, P. J. Rosenthal, J. L. DeRisi, Metagenomic next-generation sequencing of samples from pediatric febrile illness in Tororo, Uganda. PLoS One 14, e0218318 (2019).

17. E. L. Tsalik, R. Henao, M. Nichols, T. Burke, E. R. Ko, M. T. McClain, L. L. Hudson, A. Mazur, D. H. Freeman, T. Veldman, R. J. Langley, E. B. Quackenbush, S. W. Glickman, C. B. Cairns, A. K. Jaehne, E. P. Rivers, R. M. Otero, A. K. Zaas, S. F. Kingsmore, J. Lucas, V. G. Fowler, L. Carin, G. S. Ginsburg, C. W. Woods, Host gene expression classifiers diagnose acute respiratory illness etiology. Sci Transl Med 8, 322ra11 (2016).

18. A. K. Zaas, M. Chen, J. Varkey, T. Veldman, A. O. Hero, J. Lucas, Y. Huang, R. Turner, A. Gilbert, R. Lambkin-Williams, N. C. Øien, B. Nicholson, S. Kingsmore, L. Carin, C. W. Woods, G. S. Ginsburg, Gene expression signatures diagnose influenza and other symptomatic respiratory viral infections in humans. Cell Host Microbe 6, 207–17 (2009).

19. K. F. Fukutani, C. M. Nascimento-Carvalho, M. L. Bouzas, J. R. Oliveira, A. Barral, T. Dierckx, R. Khouri, H. I. Nakaya, B. B. Andrade, J. Van Weyenbergh, C. I. de Oliveira, In situ Immune Signatures and Microbial Load at the Nasopharyngeal Interface in Children With Acute Respiratory Infection. Front Microbiol 9, 2475 (2018).

20. Y.-T. Tsao, Y.-H. Tsai, W.-T. Liao, C.-J. Shen, C.-F. Shen, C.-M. Cheng, Differential Markers of Bacterial and Viral Infections in Children for Point-of-Care Testing. Trends Mol Med 26, 1118–1132 (2020).

21. L. Ashkenazi-Hoffnung, K. Oved, R. Navon, T. Friedman, O. Boico, M. Paz, G. Kronenfeld, L. Etshtein, A. Cohen, T. M. Gottlieb, E. Eden, I. Chistyakov, I. Srugo, A. Klein, S. Ashkenazi, O. Scheuerman, A host-protein signature is superior to other biomarkers for differentiating between bacterial and viral disease in patients with respiratory infection and fever without source: a prospective observational study. Eur J Clin Microbiol Infect Dis 37, 1361–1371 (2018).

22. E. Mick, A. Tsitsiklis, J. Kamm, K. L. Kalantar, S. Caldera, A. Lyden, M. Tan, A. M. Detweiler, N. Neff, C. M. Osborne, K. M. Williamson, V. Soesanto, M. Leroue, A. B. Maddux, E. A. Simões, T. C. Carpenter, B. D. Wagner, J. L. DeRisi, L. Ambroggio, P. M. Mourani, C. R. Langelier, Integrated host/microbe metagenomics enables accurate lower respiratory tract infection diagnosis in critically ill children. J Clin Invest 133 (2023), doi:10.1172/JCI165904.

23. S. M. C. Lopez, J. M. Martin, M. Johnson, M. Kurs-Lasky, W. T. Horne, C. W. Marshall, V. S. Cooper, J. V. Williams, N. Shaikh, A method of processing nasopharyngeal swabs to enable multiple testing. Pediatr Res 86, 651– 654 (2019).

24. D. E. Wood, J. Lu, B. Langmead, Improved metagenomic analysis with Kraken 2. Genome Biol 20 (2019), doi:10.1186/S13059-019-1891-0.

25. B. P. Alcock, A. R. Raphenya, T. T. Y. Lau, K. K. Tsang, M. Bouchard, A. Edalatmand, W. Huynh, A. L. V. Nguyen, A. A. Cheng, S. Liu, S. Y. Min, A. Miroshnichenko, H. K. Tran, R. E. Werfalli, J. A. Nasir, M. Oloni, D. J. Speicher, A. Florescu, B. Singh, M. Faltyn, A. Hernandez-Koutoucheva, A. N. Sharma, E. Bordeleau, A. C. Pawlowski, H. L. Zubyk, D. Dooley, E. Griffiths, F. Maguire, G. L. Winsor, R. G. Beiko, F. S. L. Brinkman, W. W. L. Hsiao, G. V. Domselaar, A. G. McArthur, CARD 2020: antibiotic resistome surveillance with the comprehensive antibiotic resistance database. Nucleic Acids Res 48, D517–D525 (2020).

26. V. A. C. de Abreu, J. Perdigão, S. Almeida, Metagenomic Approaches to Analyze Antimicrobial Resistance: An Overview. Front Genet 11, 575592 (2020).

27. E. H. Graf, K. E. Simmon, K. D. Tardif, W. Hymas, S. Flygare, K. Eilbeck, M. Yandell, R. Schlaberg, Unbiased Detection of Respiratory Viruses by Use of RNA Sequencing-Based Metagenomics: a Systematic Comparison to a Commercial PCR Panel. J Clin Microbiol 54, 1000–7 (2016).

28. F. Thorburn, S. Bennett, S. Modha, D. Murdoch, R. Gunson, P. R. Murcia, The use of next generation sequencing in the diagnosis and typing of respiratory infections. J Clin Virol 69, 96–100 (2015).

29. K. B. Waites, T. P. Atkinson, The role of Mycoplasma in upper respiratory infections. Curr Infect Dis Rep 11, 198–206 (2009).

30. F. Blasi, Clinical features of Chlamydia pneumoniae acute respiratory infection. Clin Microbiol Infect 1 **Suppl 1**, S14–S18 (1996).

31. P. J. Pérez-Chaparro, C. Gonçalves, L. C. Figueiredo, M. Faveri, E. Lobão, N. Tamashiro, P. Duarte, M. Feres, Newly identified pathogens associated with periodontitis: a systematic review. J Dent Res 93, 846–58 (2014).

32. L. Olijve, L. Jennings, T. Walls, Human Parechovirus: an Increasingly Recognized Cause of Sepsis-Like Illness in Young Infants. Clin Microbiol Rev 31 (2018), doi:10.1128/CMR.00047-17.

33. J. Zoll, S. Erkens Hulshof, K. Lanke, F. Verduyn Lunel, W. J. G. Melchers, E. Schoondermark-van de Ven, M. Roivainen, J. M. D. Galama, F. J. M. van Kuppeveld, Saffold virus, a human Theiler’s-like cardiovirus, is ubiquitous and causes infection early in life. PLoS Pathog 5, e1000416 (2009).

34. M. Venter, S. van Niekerk, A. Rakgantso, N. Bent, Identification of deletion mutant respiratory syncytial virus strains lacking most of the G protein in immunocompromised children with pneumonia in South Africa. J Virol 85, 8453–8457 (2011).

35. J. T. Sims, J. Poorbaugh, C.-Y. Chang, T. R. Holzer, L. Zhang, S. M. Engle, S. Beasley, T. N. Doman, L. Naughton, R. E. Higgs, N. Kallewaard, R. J. Benschop, Relationship between gene expression patterns from nasopharyngeal swabs and serum biomarkers in patients hospitalized with COVID-19, following treatment with the neutralizing monoclonal antibody bamlanivimab. J Transl Med 20, 134 (2022).

36. N. R. Cheemarla, A. Hanron, J. R. Fauver, J. Bishai, T. A. Watkins, A. F. Brito, D. Zhao, T. Alpert, C. B. F. Vogels, A. I. Ko, W. L. Schulz, M. L. Landry, N. D. Grubaugh, D. van Dijk, E. F. Foxman, Nasal host response-based screening for undiagnosed respiratory viruses: a pathogen surveillance and detection study. Lancet Microbe 4, e38–e46 (2023).

37. A. Wesolowska-Andersen, J. L. Everman, R. Davidson, C. Rios, R. Herrin, C. Eng, W. J. Janssen, A. H. Liu, S. S. Oh, R. Kumar, T. E. Fingerlin, J. Rodriguez-Santana, E. G. Burchard, M. A. Seibold, Dual RNA-seq reveals viral infections in asthmatic children without respiratory illness which are associated with changes in the airway transcriptome. Genome Biol 18, 12 (2017).

38. D. Butler, C. Mozsary, C. Meydan, J. Foox, J. Rosiene, A. Shaiber, D. Danko, E. Afshinnekoo, M. MacKay, F. J. Sedlazeck, N. A. Ivanov, M. Sierra, D. Pohle, M. Zietz, U. Gisladottir, V. Ramlall, E. T. Sholle, E. J. Schenck, C. D. Westover, C. Hassan, K. Ryon, B. Young, C. Bhattacharya, D. L. Ng, A. C. Granados, Y. A. Santos, V. Servellita, S. Federman, P. Ruggiero, A. Fungtammasan, C. S. Chin, N. M. Pearson, B. W. Langhorst, N. A. Tanner, Y. Kim, J. W. Reeves, T. D. Hether, S. E. Warren, M. Bailey, J. Gawrys, D. Meleshko, D. Xu, M. Couto-Rodriguez, D. Nagy-Szakal, J. Barrows, H. Wells, N. B. O’Hara, J. A. Rosenfeld, Y. Chen, P. A. D. Steel, A. J. Shemesh, J. Xiang, J. Thierry-Mieg, D. Thierry-Mieg, A. Iftner, D. Bezdan, E. Sanchez, T. R. Campion, J. Sipley, L. Cong, A. Craney, P. Velu, A. M. Melnick, S. Shapira, I. Hajirasouliha, A. Borczuk, T. Iftner, M. Salvatore, M. Loda, L. F. Westblade, M. Cushing, S. Wu, S. Levy, C. Chiu, R. E. Schwartz, N. Tatonetti, H. Rennert, M. Imielinski, C. E. Mason, Shotgun transcriptome, spatial omics, and isothermal profiling of SARS-CoV-2 infection reveals unique host responses, viral diversification, and drug interactions. Nat Commun 12 (2021), doi:10.1038/S41467-021-21361-7.

39. L. Combaret, D. Taillandier, C. Polge, D. Béchet, D. Attaix, in The Molecular Nutrition of Amino Acids and Proteins, (Elsevier, 2016), pp. 27–37.

40. P. Yu, X. Zhang, N. Liu, L. Tang, C. Peng, X. Chen, Pyroptosis: mechanisms and diseases. Signal Transduct Target Ther 6, 128 (2021).

41. S. Tristram, M. R. Jacobs, P. C. Appelbaum, Antimicrobial resistance in Haemophilus influenzae. Clin Microbiol Rev 20, 368–389 (2007).

42. J. Van Eldere, M. P. E. Slack, S. Ladhani, A. W. Cripps, Non-typeable Haemophilus influenzae, an under-recognised pathogen. Lancet Infect Dis 14, 1281–92 (2014).

43. B. Lobb, M. C. Lee, C. L. McElheny, Y. Doi, K. Yahner, A. Hoberman, J. M. Martin, J. A. Hirota, A. C. Doxey, N. Shaikh, Genomic classification and antimicrobial resistance profiling of Streptococcus pneumoniae and Haemophilus influenzae isolates associated with paediatric otitis media and upper respiratory infection. BMC Infect Dis 23, 596 (2023).

44. A. M. Saravia-Butler, J. C. Schisler, D. Taylor, A. Beheshti, D. Butler, C. Meydan, J. Foox, K. Hernandez, C. Mozsary, C. E. Mason, R. Meller, Host transcriptional responses in nasal swabs identify potential SARS-CoV-2 infection in PCR negative patients. iScience 25, 105310 (2022).

45. M. L. Landry, E. F. Foxman, Antiviral Response in the Nasopharynx Identifies Patients With Respiratory Virus Infection. J Infect Dis 217, 897–905 (2018).

46. N. R. Cheemarla, T. A. Watkins, V. T. Mihaylova, B. Wang, D. Zhao, G. Wang, M. L. Landry, E. F. Foxman, Dynamic innate immune response determines susceptibility to SARS-CoV-2 infection and early replication kinetics. J Exp Med 218 (2021), doi:10.1084/jem.20210583.

47. R. Karalus, A. Campagnari, Moraxella catarrhalis: a review of an important human mucosal pathogen. Microbes Infect 2, 547–59 (2000).

48. B. Henriques-Normark, S. Normark, Commensal pathogens, with a focus on Streptococcus pneumoniae, and interactions with the human host. Exp Cell Res 316, 1408–14 (2010).

49. S. Andrews, FastQC: a quality control tool for high throughput sequence data. (2010) (available at http://www.bioinformatics.babraham.ac.uk/projects/fastqc).

50. S. Chen, Y. Zhou, Y. Chen, J. Gu, fastp: an ultra-fast all-in-one FASTQ preprocessor. Bioinformatics 34, i884– i890 (2018).

51. P. Ewels, M. Magnusson, S. Lundin, M. Käller, MultiQC: summarize analysis results for multiple tools and samples in a single report. Bioinformatics 32, 3047–3048 (2016).

52. S. F. Altschul, T. L. Madden, A. A. Schäffer, J. Zhang, Z. Zhang, W. Miller, D. J. Lipman, Gapped BLAST and PSI-BLAST: A new generation of protein database search programs. Nucleic Acids Res 25, 3389–3402 (1997).

53. B. D. Ondov, T. J. Treangen, P. Melsted, A. B. Mallonee, N. H. Bergman, S. Koren, A. M. Phillippy, Mash: fast genome and metagenome distance estimation using MinHash. Genome Biol 17, 132 (2016).

54. P. Petrenko, B. Lobb, D. A. Kurtz, J. D. Neufeld, A. C. Doxey, MetAnnotate: function-specific taxonomic profiling and comparison of metagenomes. BMC Biol 13, 92 (2015).

55. H. Wickham, ggplot2: Elegant Graphics for Data Analysis (Springer-Verlag, New York, 2016).

56. A. Bankevich, S. Nurk, D. Antipov, A. A. Gurevich, M. Dvorkin, A. S. Kulikov, V. M. Lesin, S. I. Nikolenko, S. Pham, A. D. Prjibelski, A. V Pyshkin, A. V Sirotkin, N. Vyahhi, G. Tesler, M. A. Alekseyev, P. A. Pevzner, SPAdes: a new genome assembly algorithm and its applications to single-cell sequencing. J Comput Biol 19, 455–77 (2012).

57. B. Bushnell, BBMap: A Fast, Accurate, Splice-Aware Aligner https://www.osti.gov/biblio/1241166 (2014).

58. R. C. Edgar, MUSCLE: multiple sequence alignment with high accuracy and high throughput. Nucleic Acids Res 32, 1792–7 (2004).

59. M. Price, P. Dehal, A. Arkin, FastTree 2--approximately maximum-likelihood trees for large alignments. PLoS One 5 (2010), doi:10.1371/JOURNAL.PONE.0009490.

60. R. Patro, G. Duggal, M. I. Love, R. A. Irizarry, C. Kingsford, Salmon provides fast and bias-aware quantification of transcript expression. Nat Methods 14, 417–419 (2017).

61. M. I. Love, W. Huber, S. Anders, Moderated estimation of fold change and dispersion for RNA-seq data with DESeq2. Genome Biol 15 (2014), doi:10.1186/S13059-014-0550-8.

62. M. V. Kuleshov, M. R. Jones, A. D. Rouillard, N. F. Fernandez, Q. Duan, Z. Wang, S. Koplev, S. L. Jenkins, K. M. Jagodnik, A. Lachmann, M. G. McDermott, C. D. Monteiro, G. W. Gundersen, A. Maayan, Enrichr: a comprehensive gene set enrichment analysis web server 2016 update. Nucleic Acids Res 44, W90–W97 (2016).

